# More than privacy: Australians’ concerns and misconceptions about the COVIDSafe App

**DOI:** 10.1101/2020.06.09.20126110

**Authors:** Rae Thomas, Zoe A Michaleff, Hannah Greenwood, Eman Abukmail, Paul Glasziou

## Abstract

**Background:** Timely and effective contact tracing is an essential public health role to curb the transmission of COVID-19. App-based contact tracing has the potential to optimise the resources of overstretched public health departments. However, it’s efficiency is dependent on wide-spread adoption. We aimed to identify the proportion of people who had downloaded the Australian Government COVIDSafe app and examine the reasons why some did not.

**Method:** An online national survey with representative quotas for age and gender was conducted between May 8 and May 11 2020. Participants were excluded if they were a healthcare professional or had been tested for COVID-19.

**Results:** Of the 1802 potential participants contacted, 289 were excluded, 13 declined, and 1500 participated in the survey (response rate 83%). Of survey participants, 37% had downloaded the COVIDSafe app, 19% intended to, 28% refused, and 16% were undecided. Equally proportioned reasons for not downloading the app included privacy (25%) and technical concerns (24%). Other reasons included a belief that social distancing was sufficient and the app is unnecessary (16%), distrust in the Government (11%), and apathy (11%). In addition, COVIDSafe knowledge varied with confusion about its purpose and capabilities.

**Conclusion:** For the COVIDSafe app to be accepted by the public and used correctly, public health messages need to address the concerns of its citizens, specifically in regards to privacy, data storage, and technical capabilities. Understanding the specific barriers preventing the uptake of tracing apps provides the opportunity to design targeted communication strategies aimed at strengthening public health initiatives such as download and correct use.

## Introduction

In the absence of a vaccine, non-drug interventions to prevent COVID-19 and any other future infectious outbreaks, are critical [1,2]. The public have been asked to practice preventive behaviours such as hand hygiene, physical distancing, quarantining and getting tested when sick. These behaviours are being promoted by national and international public health organisations through population-based communication strategies. Alongside individually practiced prevention strategies, are population-based strategies such as contact tracing, which is also critical to prevent or slow the spread of disease. To improve public health tracing and the speed at which this occurs, several countries have introduced app-based contact tracing. Contact tracing apps vary in design from reporting symptoms to public health authorities [3] to allowing access to phone data after testing positive to COVID-19 [4] and in whether the data is centralized [5]. Those apps in current use have had varying degrees of success [3,6] Since the Australian Government launched the COVIDSafe app [4] over 6 million (almost 25%) Australians have downloaded the app, but this is still short of the 40% proposed target to be effective. Worldwide concerns have been raised about privacy and ethics of this digital approach [7] which may hamper downloads and decrease effectiveness.

App-based contact tracing requires public cooperation. Individuals are required to install the app, keep bluetooth functions on, have the app activated or open on their phone, and carry the phone with them when outside of their home. This sounds simple, but when considered from a behaviour change perspective, these behaviours are complex and need to be performed together to optimise contact tracing functionality [8]. To identify behaviour change techniques to improve the uptake of app-based tracing, we first need to understand people’s reasons for not downloading the app. In this study, we aimed to identify Australian’s understanding of the purpose and capabilities of the Australian Government’s COVIDSafe app and explore reasons why some Australians chose not to download the app.

## Method

Participants were recruited for a national, cross-sectional, online survey by a panel provider (Dynata). The sample was representative of all Australian states and territories with quotas for age and gender. Participants were included if aged 18 years or over, and excluded if they had, or thought they had COVID-19, or were healthcare professionals.

All participants were asked whether they had, or intended to download the COVIDSafe app. If they responded “unsure” or “no intention to download”, they were asked to provide a reason for their response. We qualitatively coded the reasons for inaction and uncertainty. Participants then rated their strength of agreement on six statements related to the app’s purpose and capability using a 5-point Likert scale (1=strongly disagree to 5=strongly agree and a don’t know response) The survey items and response scale is available in the Supplementary file.

## Results

Of the 1802 potential participants contacted, 289 were excluded, 13 declined and 1500 participated in the survey (response rate 83%). There was a representation across adult age groups and gender (50% male), and education levels were distributed evenly (highschool and TAFE qualification or lower 49%; tertiary qualification 51% (see Table).

**Table 1.** Participant characteristics (N=1500)

| **Characteristics** | **N** | **%** |
| --- | --- | --- |
| Female | 750 | 50 |
| Age |  |  |
| 18-24 y | 171 | 11 |
| 25-34 y | 264 | 18 |
| 35-44 y | 239 | 16 |
| 45-54 y | 223 | 15 |
| 55-64 y | 222 | 15 |
| 65-74 y | 227 | 15 |
| 75 or older | 154 | 10 |
| Education |  |  |
| High school graduate or Less | 459 | 31 |
| Trade Certificate (I-IV) | 276 | 18 |
| Tertiary | 765 | 51 |
| Australian States and Territories |  |  |
| QLD | 302 | 20 |
| NSW | 471 | 31 |
| ACT | 29 | 2 |
| NT | 9 | 1 |
| WA | 160 | 11 |
| VIC | 382 | 25 |
| TAS | 34 | 2 |
| SA | 113 | 7 |
| Aboriginal or Torres Strait Islander |  |  |
| Yes | 17 | 1 |
| No | 1471 | 98 |
| Prefer not to say | 12 | 1 |
| Born in Australia | 1049 | 70 |

Of survey participants, 37% (555/1500) said they had downloaded the COVIDSafe app, 19% (285/1500) had intended to, 28% (420/1500) refused, and 16% (240/1500) were undecided. Of those who refused or were undecided (n=660), 25% cited privacy concerns as their primary reason. For example, many distrusted the security of the app: that is the COVIDSafe app wasn’t safe and it could be hacked and their information used without authority. Importantly, another 24% cited technical problems such as: their phone was too old or had data consumption and storage space limitations. Other reasons included a belief that social distancing was sufficient and the app is unnecessary for them (16%), distrust in the Government (11%), questioning the app’s effectiveness (7%), wanting to explore more information before deciding (5%), and other miscellaneous responses (11%) including apathy and following the decisions of others.

With respect to the app’s intended purpose and capabilities most participants correctly agreed that the app would make contact tracing faster and easier (75%) and that more potentially exposed people would be found and informed (73%; see Figure). In contrast, 50% of participants thought their personal information would be shared post-pandemic and incorrectly thought the app would detect when people with COVID-19 were near them (76%). Interestingly, participants were almost divided in knowing whether the app would inform them it was safe to leave their house (see Figure).

**Figure.**
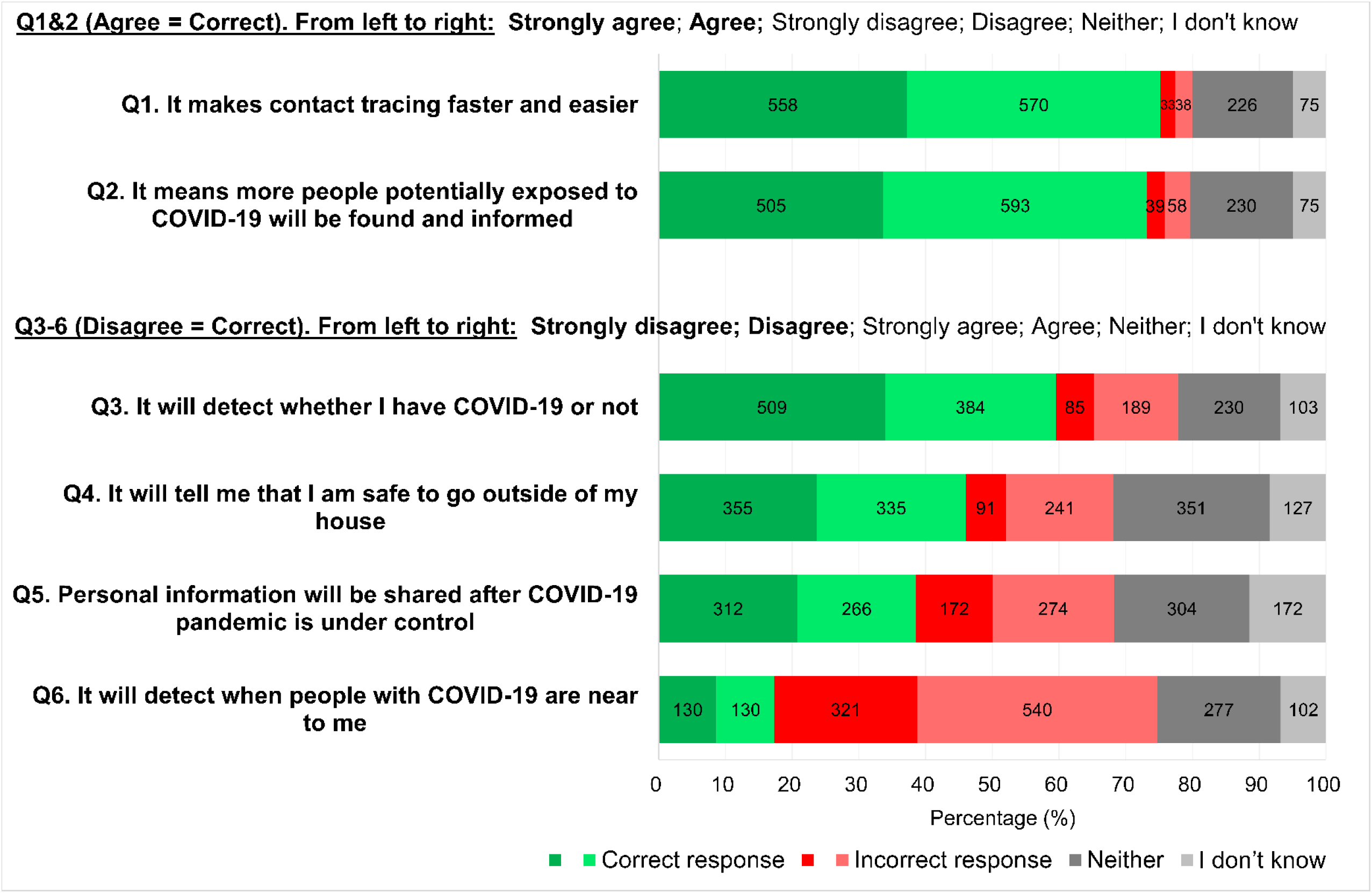
Participants’ ratings of suggested purposes and capabilities of the COVIDSafe app (N=1500).

## Discussion

Timely and effective contact tracing is an essential public health measure to curb the transmission of COVID-19. Contact tracing apps are controversial in their design and level of effectiveness [3,5,6] but they might have the potential to prevent wide-spread community transmission and optimise the resources of overstretched public health organisations [9]. An important driver in it’s efficiency is wide-spread public adoption [9]. Our study identified Australian participants’ understanding about Australian Government’s COVIDSafe app and explored reasons why some Australians chose not to use it.

Approximately 37% of our sample said they had downloaded the app and 19% intended to. This proportion is concordant with another Australian survey with a smaller online sample (n=500), where 44% of participants reported downloading the COVIDSafe app [10]. However, acceptability of tracing apps were higher in other countries when participants were asked whether they intended to download hypothetical apps. For example, in a recent online survey from Ireland (N>8000) [11], when asked about downloading a tracing app not yet available, 58% of participants said they would download it and 25% said they probably would [8]. While in another online survey with almost 6000 participants from five countries (UK, Germany, Italy, France and the US), 75% of participants said they definitely or probably would install a tracing app [12]. It will be interesting to see whether people’s intention is actioned by a subsequent download of a contact tracing app.

In open-ended text responses, 25% of participants in our study who reported inaction in downloading the COVIDSafe app were concerned about privacy. This is lower than the 31% in the smaller Australian study who identified privacy as a problem [10] and 41% of participants in the Irish study [11]. These differences may be due to free text responses available in our survey compared with listed option responses in the other studies. Additionally, compared with only 11% of participants in our survey, there was more distrust in government surveillance post-pandemic in Irish participants (33%) [11], and with participants in the cross-country survey (42%) [12]. It appears that when considering communication strategies to improve contact tracing app downloads and use, better communication approaches are warranted to put the public’s privacy concerns and government distrust at ease.

Our study also demonstrates missed communication opportunities to correct erroneous beliefs about the capabilities of the COVIDSafe. Over half of our participants thought the COVIDSafe app would or might tell them when it was safe to leave the house and 40% thought it would or might tell them whether they had COVID-19. Addressing these perceptions and issues about the capabilities of the app in public messaging will be important to achieving sufficient uptake of tracing apps.

To our knowledge, this study is the first to qualitatively analyse open-ended text responses to barriers for downloading a contact tracing app. This approach (rather than listed options) decreases potential researcher biases and strengthens the ability to inform communication techniques targeted to improve app uptake. However, we didn’t assess cultural and linguistic diversity which limits the generalisability of our findings and these results may not reflect the perceptions of individuals where English is not their primary language.

Although, their level of effectiveness in contact tracing is still unclear [3,6], apps have been used in various countries to help with both contact tracing [4] and with medical management of COVID-19 cases [13]. Their utility lies as an adjunct tracing strategy alongside public health staff, particularly when community transmissions are high. For apps to be accepted by the public and used correctly, we need to better communicate concerns about privacy, data storage, and technical capabilities. Lessons learned in this pandemic will be invaluable for future, inevitable infectious outbreaks.

## Data Availability

Data may be available upon request from authors.

## Acknowledgements

RT, ZAM, and HL are supported by a National Health and Medical Research Council Program grant (#1106452). PG is supported by a NHMRC Research Fellowship (#1080042).

## Supplementary File

**Supplementary Table.** COVIDSafe survey items and scores

|  |  |
| --- | --- |
| • It makes contact tracing faster and easier | 1 – 5 strongly disagree to strongly agree and a 'don't know' option |
| • It means more people potentially exposed to COVID-19 will be found and informed | 1 – 5 strongly disagree to strongly agree and a 'don't know' option |
| • It makes it easier to track contacts across state borders | 1 – 5 strongly disagree to strongly agree and a 'don't know' option |
| • Personal information will be shared after COVID-19 pandemic is under control | 1 – 5 strongly disagree to strongly agree and a 'don't know' option |
| • It will tell me that I am safe to go outside of my house | 1 – 5 strongly disagree to strongly agree and a 'don't know' option |
| • It will detect when people with COVID-19 are near to me | 1 – 5 strongly disagree to strongly agree and a 'don't know' option |
| • It will detect whether I have COVID-19 or not | 1 – 5 strongly disagree to strongly agree and a 'don't know' option |

## References

1. Hoffmann T, Glasziou P. What if the vaccine or drugs don’t save us? Plan B for coronavirus means research on alternatives is urgently needed. Online: The Conversation; 2020. Available from: https://theconversation.com/what-if-the-vaccine-or-drugs-dont-save-us-plan-b-for-coronavirus-means-research-on-alternatives-is-urgently-needed-136833 Accessed July 2020

2. Michie S, West R. Behavioural, environmental, social, and systems interventions against COVID-19. BMJ. 2020;370:m2982

3. Kendall M, Milsom L, Abeler-DÖrner L, Wymant C, Ferretti L, Briers M et al. COVID-19 incidence and R decreased on the Isle of Wight after launch of the Test, Trace, Isolate programme. medRxiv doi:10.1101/2020.05.05.20091587

4. Australian Government, Department of Health. COVIDSafe app. Available from: https://www.health.gov.au/resources/apps-and-tools/covidsafe-app Accessed May 2020

5. Leins K, Culnane C, Rubinstein BIP. Tracking, tracing, trust: Contemplating mitigating the impact of COVID-19 through technological interventions. Med J Aust 2020;213(1):6- 8.e1. doi:10.5694/mja2.50669

6. Wise J. COVID-19: test and trace system is not fit for purpose, says Independent SAGE. BMJ 2020;369:m2302

7. Abeler J, Bäcker M, Buermeyer U, Zillessen H. COVID-19 Contact tracing and data protection can go together. JMIR Mhealth Uhealth 2020;8(4):e19359) doi: 10.2196/19359

8. Michie S, Atkins L, West R. The behaviour change wheel. A guide to designing interventions. 1st ed. Great Britain: Silverback Publishing. 2014:1003–10.

9. Currie DJ, Peng CQ, Lyle DM, Jameson BA, Frommer MS. Stemming the flow: how much can the Australian smartphone app help to control COVID-19? Public Health Res Pract. 2020;30(2):e3022009

10. Garret P, White J, Little D, et al. A representative sample of Australian participant’s attitudes towards the COVIDSafe App. Available from: https://paulgarrettphd.github.io/Site/Wave3PrelimAnalysis.html Accessed May 2020

11. O’Callaghan ME, Buckley J, Fitzgerald B, Johnson K, Laffey J, McNicholas B, et al. A national survey of attitudes to COVID-19 digital contact tracing in the Republic of Ireland. Research Square doi:10.21203/rs.3.rs-40778/v1

12. Altmann S, Milsom L, Zillessen H, Blasone R, Gerdon F, Bach R, et al. Acceptability of app-based contact tracing for COVID-19: cross-country survey evidence. medRxiv doi:10.1101/2020.05.05.20091587

13. Echeverria P, Bergas MAM, Puig J, Isnard M, Massot M, Vedia C, et al. COVIDApp as an innovative strategy for the management and follow-up of COVID-19 cases in long-term facilities in Catalonia: Implementation study. JMIR Public Health Surveillance. 2020:6(3);e21163

